# Metformin associates with improved myocardial perfusion reserve and survival in patients with Type 2 Diabetes Mellitus

**DOI:** 10.1101/2023.04.13.23288441

**Authors:** Noor Sharrack, Kristopher D Knott, Gaurav S Gulsin, Tushar Kotecha, Louise AE Brown, Jian L Yeo, Aldostefano Porcari, Robert D Adam, Sharmaine Thirunavukarasu, Amrit Chowdhary, Eylem Levelt, James C Moon, Gerry P McCann, Marianna Fontana, Peter Kellman, Theresa Munyombwe, Christopher Gale, David L Buckley, John P Greenwood, Peter P Swoboda, Sven Plein

## Abstract

**Background:** Metformin is an antihyperglycemic agent frequently used in the treatment of Type 2 Diabetes Mellitus (T2DM). Patients with T2DM are at increased risk of cardiovascular diseases, including coronary artery disease (CAD), silent myocardial infarction (MI) and coronary microvascular dysfunction (CMD), all of which can be detected and quantified using Cardiovascular Magnetic Resonance (CMR). We explored the association between metformin use, stress Myocardial Blood Flow (MBF), Myocardial Perfusion Reserve (MPR), survival and major adverse cardiovascular and cerbrovasular events (MACCE; a composite of all-cause death, MI, stroke, heart failure hospitalisation and coronary revascularisation) in patients with T2DM.

**Methods:** A multi-centre study of patients with T2DM, and a cohort of healthy controls underwent quantitative myocardial perfusion CMR. Global MBF and MPR were derived using an automatic artificial intelligence-supported process. Multivariable regression analysis and cox proportional hazard models quantified associations between metformin use, MBF, MPR, all-cause death and MACCE.

**Results:** Analysis included 572 patients with T2DM (68% prescribed metformin) with median follow-up 851 days (interquartile range 935-765). Metformin use was associated with an increase in MPR of 0.12 [0.08-0.40], P=0.004. There was a total of 82 (14.3%) first MACCE in all T2DM patients including a total of 25 (4.4%) deaths. Although the number of first MACCE events was similar for patients prescribed metformin (53 (14%)) compared to those who were not (29 (15.8%) (P=0.73)), there was a total of 9 deaths (2.3%) in patients prescribed metformin compred to 16 (8.7%) in patients who were not, adjusted hazard ratio 0.29 [95% CI 0.12-0.73] P=0.009).

**Conclusion:** In patients with T2DM, metformin use is associated with higher MPR and improved survival.

**Clinical Perspective:** Patients with Type 2 Diabetes Mellitus (T2DM) are at increased risk of cardiovascular disease. Cardiovascular Magnetic Resonane (CMR) can be used to detect and quantify absolute stress myocardial blood flow (MBF) and myocardial perfusion reserve (MPR), both of which are objective measures of coronary microvascular function. Metformin is frequently used in the treatment of T2DM. We investigated the association between metformin use, CMR-derived stress MBF, MPR and clinical outcomes in patients with T2DM.

In a longitudinal cohort study of patients with T2DM, metformin use was associated with higher MPR as a marker of microvascular function, and improved survival after adjusting for certain confounding parameters.

Further prospective studies are needed to confirm the association between metformin use and improved MPR and reduced mortality, as well as to clarify the mechanisms responsible and quantify the dose these associated outcomes are observed.

**Central illustration:** 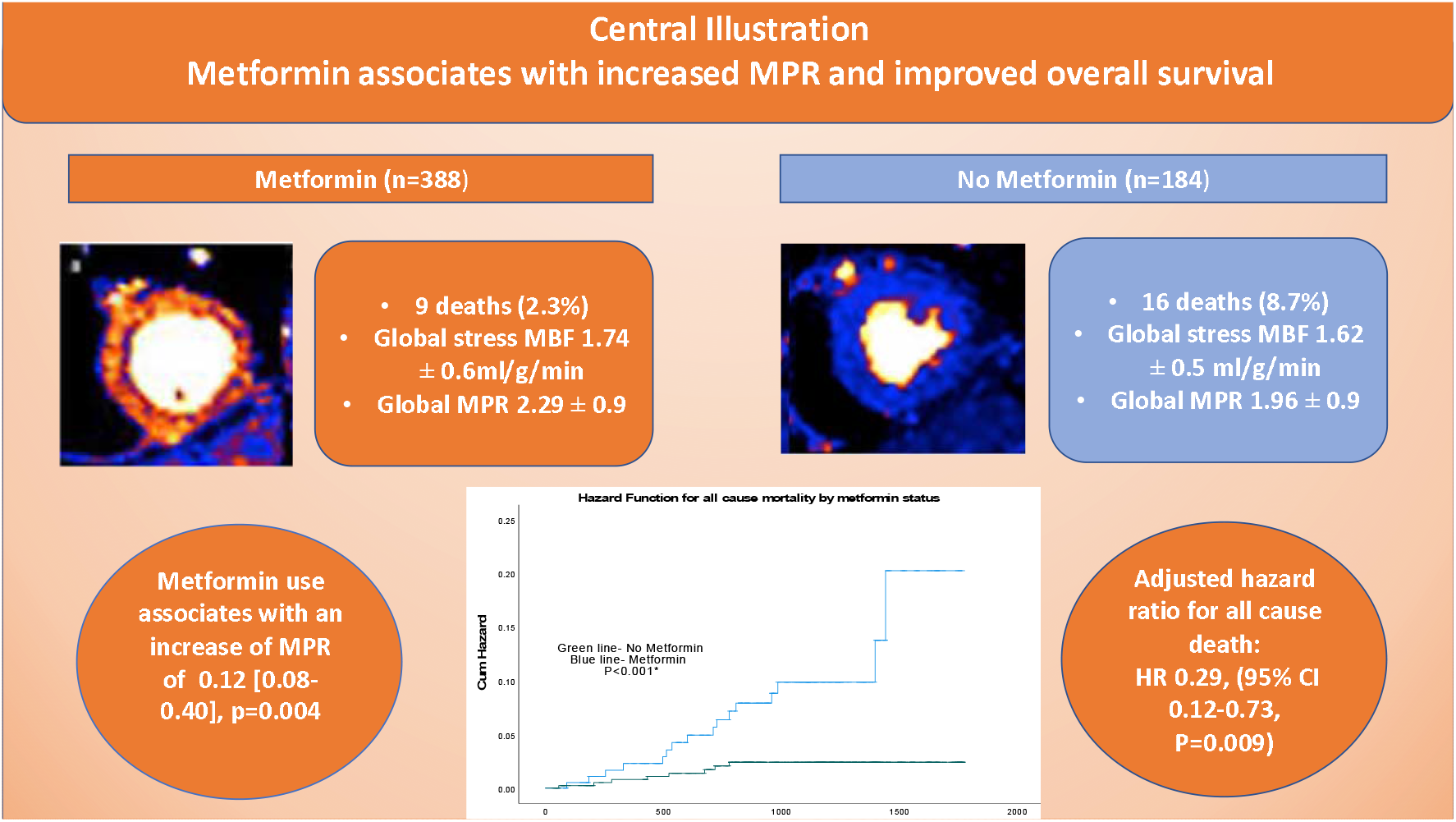

## Introduction

Cardiovascular disease is the commonest cause of death in patients with Type 2 Diabetes Mellitus (T2DM)^1^. T2DM is associated with an increased risk of epicardial coronary artery disease (CAD), silent myocardial infarction (MI) and coronary microvascular dysfunction (CMD)^2-4^.

Metformin is the most widely used antihyperglycemic agent worldwide and is first-line in most recommendations for the treatment of patients with T2DM^5-8^. As well as its glucose lowering and weight loss benefits), it was also shown to be associated with a 39% lower risk of myocardial infarction compared to conventional therapy in the United Kingdom Prospective Diabetes (UKPDS) Study^9^. In a more recent study, metformin use was associated with a nearly 40% reduction in cardiovascular mortality when compared to sulfonylurea monotherapy^10^.

Metformin may improve endothelial function as well as reducing insulin resistance in patients with T2DM^11, 12^. In non-diabetic women with chest pain and angiographically normal coronary arteries, metformin was shown to improve coronary microvascular function and decrease MI incidence^13^. In the retrospective Prevention of REStenosis with Tranilast and its Outcomes (PRESTO) trial, the use of metformin in diabetic patients undergoing coronary intervention decreased the risk of death and MI (relative risk reduction of 79%) compared to patients treated with a sulfonylurea or insulin^14^. Nevertheless, the mechanism of benefit of metformin is not clear and warrants further investigation, since meta-analyses of intensive versus normal glucose control have shown minimal benefit on cardiovascular outcomes^15^.

CMR allows simultaneous assessment of cardiac function, detection of MI, regional ischaemia suggesting epicardial CAD and, with quantitative perfusion analysis, absolute measures of myocardial blood flow (MBF) for the assessment of coronary microvascular dysfunction. Previous studies have shown that patients with T2DM have reduced global myocardial perfusion reserve (MPR) and stress MBF, independent of significant flow limiting epicardial CAD, suggestive of CMD^16, 17^. We aimed to test the hypothesis that metformin use in patients with T2DM is associated with improved stress MBF and MPR and is associated with improved survival and reduced major adverse cardiovascular and cerbrovasular events (MACCE; a composite of all-cause mortality, MI, stroke, heart failure hospitalisation and coronary revascularisation).

## Methods

### Participants

Patients aged 18 years or older, with a diagnosis of T2DM, undergoing quantitative stress perfusion CMR, were recruited from four United Kingdom centres (St Bartholomew’s Hospital, Barts Heart Centre, London; Glenfield Hospital, University Hospitals of Leicester NHS Trust, Leicester; Leeds Teaching Hospitals NHS Foundation Trust, Leeds and the Royal Free Hospital, Royal Free London NHS Foundation Trust, London) between September 2016 and May 2021. Diagnosis of T2DM was based on a HbA1c >48mmol/l or a known diagnosis of T2DM. Exclusion criteria included contraindications to adenosine, gadolinium-based contrast or MRI.

All participants had received quantitative myocardial perfusion stress CMR for clinical or research indications. Those with sub-optimal image quality, mistriggering or other artefacts were excluded.

### Controls

We recruited 52 age- and sex-matched, healthy controls with no past medical history of cardiac disease or major risk factors from two cardiac centres (Leeds Teaching Hospitals and Glenfield Hospital) between September 2016 and May 2021. Exclusion criteria included a known past medical history of hypertension, hypercholesterolaemia, diabetes mellitus (DM), smoking, previous CAD or revascularisation or perfusion defect on stress CMR, contraindications to adenosine, gadolinium-based contrast or MRI and subsequent evidence of abnormal late gadolinium enhancement (LGE) on CMR.

### Clinical Outcomes

Patient co-morbidities and clinical outcomes were collated from electronic patient records and the National Health Service (NHS) Spine Portal through deterministic linkage using each patient’s unique NHS identification number. Co-morbidities recorded included hypertension, dyslipidaemia, atrial fibrillation, CAD, coronary revascularisation (percutaneous coronary intervention (PCI) or coronary artery bypass graft (CABG) surgery, MI, stroke or transient ischaemic attack (TIA) and cancer. All patients had provided written, informed consent and had more than one year of follow-up data available.

The joint primary outcomes were all-cause mortality and MACCE (a composite of all-cause mortality, MI, stroke, heart failure hospitalisation and coronary revascularisation). Where patients had more than one MACCE, the first event was used for analysis.

### Cardiovascular Magnetic Resonance imaging

CMR scans were performed on 1.5T (Magnetom Aera, Siemens Healthcare, Erlangen, Germany) or 3T (Prisma, Siemens Healthcare, Erlangen, Germany) scanners with a standard protocol consisting of localisers, short-axis and long-axis cine imaging, perfusion imaging and late gadolinium enhancement (LGE) imaging. 169 (30%) patients were scanned on 1.5T and 403 (70%) patients were scanned on 3T. All healthy controls were scanned on 3T. Perfusion imaging on 1.5T scanners used a steady state free precession (SSFP) pulse seqeunce and on 3T scanners a fast single-shot gradient echo (FLASH) pulse sequence. All participants were instructed to abstain from caffeine for 24 hours before the study. For perfusion imaging, adenosine was infused for a minimum of 3 minutes, at a rate of 140 micrograms/kg/min and increased up to a maximum of 210 micrograms/kg/min if there was insufficient haemodynamic response (heart rate increase less than 10bpm) or there was no symptomatic response, in line with standard clinical practice guidelines^18^. Images were acquired during free breathing over at least 60 dynamics to allow for any patients with poor ventricular function and consequent slow contrast uptake. A minimum interval duration of ten minutes was kept between stress and subsequent rest perfusion acquisitions. Participants’ blood pressures and heart rates were recorded at regular intervals during adenosine infusion. For each perfusion acquisition, an intravenous bolus of (0.05-0.75mmol/kg) gadobutrol (Gadovist, Leverkusen, Germany) or gadoterate meglumine (Dotarem, Guerbet, Paris, France) was administered at 4-5ml/s. Perfusion mapping was performed in three short axis sections, using a dual sequence technique combining a low-resolution arterial input function acquisition and a higher resolution myocardial perfusion acquisition as previously described by Kellman et. al^19^.

### Image Analysis

Image analysis was undertaken by experienced operators at the site core lab. Those with poor image quality were excluded. Measurement of cardiac volume parameters and the presence of LGE were made using cvi42 software (Circle Cardiovascular Imaging, Calgary, Canada). Left ventricular systolic and diastolic volume, ejection fraction, and the presence and distribution (ischaemic or non-ischaemic) of LGE were recorded. Non-ischaemic LGE refers to late gadolinium enhancement in a mid-myocardial or subepicardiaI pattern, as opposed to ischaemic LGE, which is subendocardial. Rest and stress perfusion maps were generated inline by an automatic artificial intelligence-supported process whereby myocardial blood flow is quantified for each pixel of the myocardium in ml/g/min. By averaging all pixel values in the 3 slices, global MBF and global MPR (ratio of stress to rest MBF) were derived^19^. Automatically derived perfusion maps were reviewed by experienced observers to exclude data affected by gating and motion correction artefact and partial voluming affects.

### Statistical Analysis

Statistical analysis was performed using SPSS (IBM SPSS Statistics, version 27.0). Normality was assessed through the Shaprio-Wilk test and variance was assessed by the Leven’s test for equality of variance. Continuous variables are presented as mean±SD. Categorical variables are presented as absolute values and percentages. Means were compared using the appropriate test, student *t* test for continuous variables and chi-squared (χ^2^) test for categorical variables. A *P* value of <0.05 was considered statistically significant.

Multivariable linear regression analysis, adjusting for age, sex, left ventricular ejection fraction (LVEF), left ventricular mass (LV mass), left ventricular end diastolic volume (LVEDV), hypertension, evidence of previous PCI, CABG surgery, MI, body surface area (BSA) and ethnicity was undertaken to seek association between metformin, stress MBF and MPR. Cox proportional hazard models adjusting for comorbidities and CMR parameters sought associations between stress MBF and MPR with all-cause mortality and MACCE. For the all-cause mortality model, we adjusted for age and LVEF (only two predictors using the 10 events per parameter rule of thumb) and for the MACCE model we adjusted for 8 parameters including age, LVEF, sex, LVEDV, LV mass, stress MBF, MPR and hypertension. Kaplan-Meier curves represented hazard curves. Patient data was categorised depending on whether the patient was prescribed metformin at the time of the CMR scan.

## Results

### Patient group

A total of 572 patients with T2DM were included with a median follow-up of 851 days (IQR 935-765) days.

#### Patient characteristics

Patient characteristics are shown in table 1 and are presented according to whether subjects were prescribed metformin at the time of their CMR. Their mean age was 65± 10 years and their mean HbA1c was 59± 16 mmol/mol. 25% of all patients (n=142) had known CAD with previous MI, PCI or CABG surgery. 68% of all patients (n=388) were prescribed metformin. Metformin positive and Metformin negative groups were well-matched for age and sex. Patients prescribed metformin were more likely to have had previous PCI, MI or CABG surgery (28% vs 19%, P=0.027) compared to those not prescribed metformin. They were also more likely to be of Asian ethnicity (32% vs 22%, p=0.01), suffer from hypercholesterolaemia (62% vs 54%, P=0.044) and have peripheral vascular disease (10% vs 4%, P=0.016). Patients who were prescribed metformin had higher mean HbA1c levels than those not prescribed metformin (60±16 mmol/mol vs 56±15 mmol/mol, respectively, P=0.022). However, patients not prescribed metformin had a higher prevelance of atrial fibrillation (21% vs 10%, P<0.001), previous stroke or transient ischaemic attack (10% vs 4%, P= 0.008) and history of current smoking (25% vs 22%, p<0.001) compared to the non-metformin cohort.

**Table 1:**
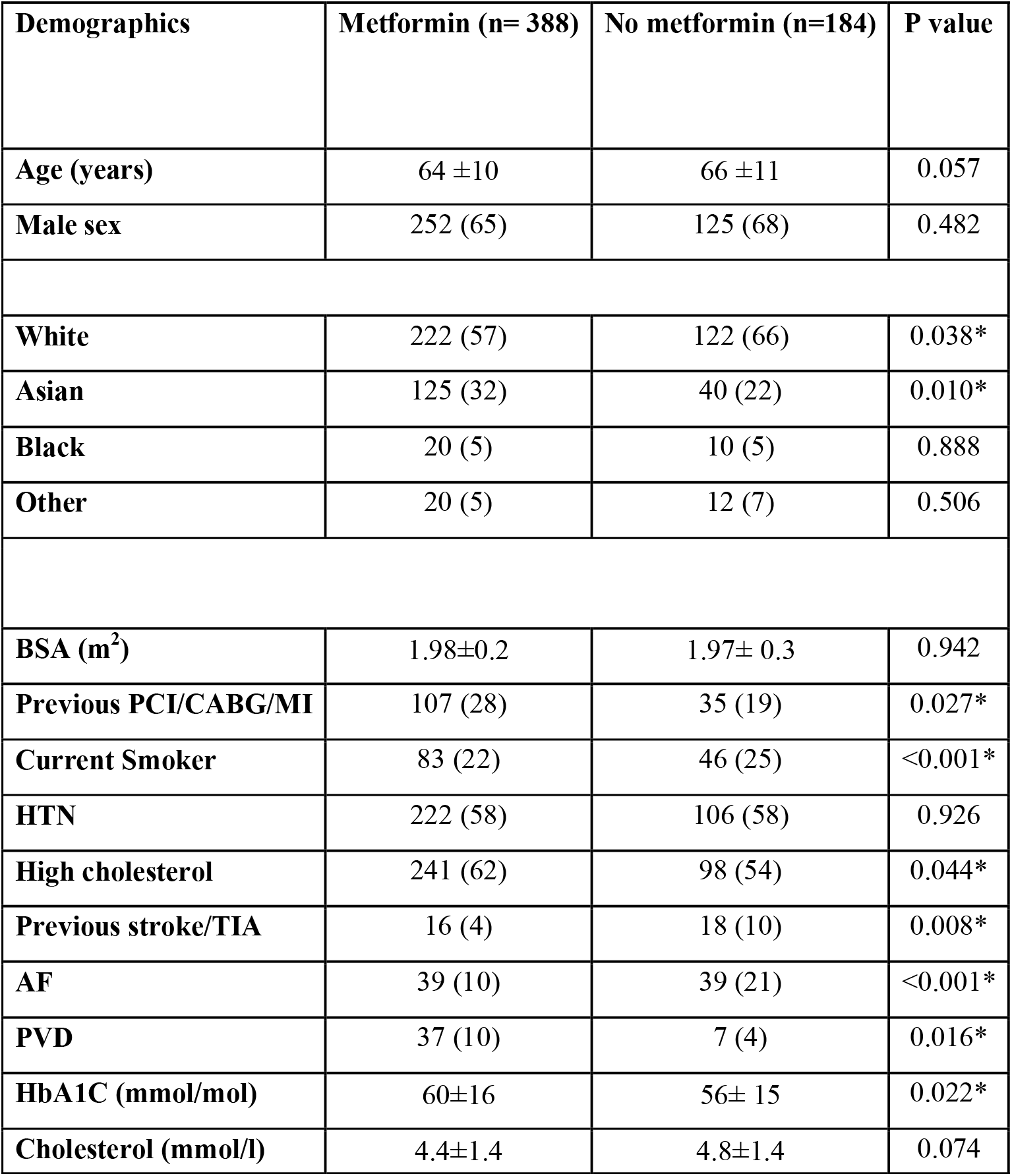
Patient demographics by metformin status. P value is considered significant at the <0.05 and indicated by*. Continuous variables are presented as mean ± SD. Dichotomous variables are presented as number (%). BSA: Body Surface Area, PCI: Percutaneous Coronary Intervention, CABG: Coronary Artery Bypass Graft surgery, MI: Myocardial Infarction, HTN: Hypertension, TI: Transient Ischaemic Attack, AF: Atrial Fibrillation, PVD: Peripheral Vascular Disease.

| Demographics | Metformin (n= 388) | No metformin (n=184) | P value |
| --- | --- | --- | --- |
| Age (years) | 64 ±10 | 66 ±11 | 0.057 |
| Male sex | 252 (65) | 125 (68) | 0.482 |
| White | 222 (57) | 122 (66) | 0.038* |
| Asian | 125 (32) | 40 (22) | 0.010* |
| Black | 20 (5) | 10 (5) | 0.888 |
| Other | 20 (5) | 12 (7) | 0.506 |
| BSA (m<sup>2</sup>) | 1.98±0.2 | 1.97± 0.3 | 0.942 |
| Previous PCI/CABG/MI | 107 (28) | 35 (19) | 0.027* |
| Current Smoker | 83 (22) | 46 (25) | <0.001* |
| HTN | 222 (58) | 106 (58) | 0.926 |
| High cholesterol | 241 (62) | 98 (54) | 0.044* |
| Previous stroke/TIA | 16 (4) | 18 (10) | 0.008* |
| AF | 39 (10) | 39 (21) | <0.001* |
| PVD | 37 (10) | 7 (4) | 0.016* |
| HbA1C (mmol/mol) | 60±16 | 56± 15 | 0.022* |
| Cholesterol (mmol/l) | 4.4±1.4 | 4.8±1.4 | 0.074 |

Patients prescribed metformin were more likely to be on antiplatelet medication, statins, gliclazide, sodium glucose cotransporter 2 (SGLT-2) inhibitors, glucagon-like peptide 1 (GLP-1) agonists and calcium channel blockers (CCBs) (table 2). However, patients not prescribed metformin, were more likely to be on mineralocorticoid receptor antagonists (MRAs), diuretics or anticoagulants.

**Table 2:** Patient medication divided by metformin status. P value is considered significant at the <0.05 level and indicated by*. Dichotomous variables are presented as number (%). DOAC: Direct Oral AntiCoagulant; ACE: Angiotensin Converting Enzyme Inhibitor; ARB: Angiotensin Receptor Blocker; MRA: Mineralocorticoid Receptor Antagonist; CCB: Calcium Channel Blocker; SGLT-2: Sodium Glucose Cotransporter-2 inhibitor; GLP-1: Glucago-Like Peptide 1 agonist.

| Medication | Metformin<br>(n=388) | No Metformin<br>(n=184) | P value |
| --- | --- | --- | --- |
| **Aspirin/ other antiplatelet** | 254 (65) | 89 (48) | <0.001* |
| **DOAC/ Warfarin** | 41 (11) | 34 (18) | 0.012* |
| **Statin** | 332 (86) | 122 (66) | <0.001* |
| **Beta blocker** | 209 (54) | 114 (62) | 0.086 |
| **ACE/ARB/Entresto** | 266 (69) | 134 (72) | 0.478 |
| **MRA** | 29 (8) | 35 (19) | <0.001* |
| **Diuretics** | 87 (22) | 68 (37) | <0.001* |
| **CCB** | 116 (30) | 40 (22) | 0.034* |
| **Nitrates** | 54 (14) | 21 (11) | 0.382 |
| **Insulin** | 53 (14) | 33 (18) | 0.199 |
| **Gliclazide** | 94 (23) | 17 (9) | <0.001* |
| **SGLT-2** | 44 (11) | 9 (5) | 0.037* |
| **GLP-1** | 61 (16) | 15 (8) | 0.011* |
| **Gliptin** | 25 (6) | 5 (3) | 0.059 |

### Healthy control group

Subjects in the healthy control group were not taking any medication. The mean age was 64±6 years and 65% of them were of male sex.

#### CMR analysis

### Patient group

169 (29.5%) of patients were scanned on 1.5T scanners and 403 (70.5%) were scanned on 3T scanners. The mean ejection fraction of diabetic patients was 57±15% (table 3). 164 (29%) had evidence of ischaemic scar on LGE indicating previous MI (table 3). Mean global stress MBF in all patients was 1.70± 0.6 ml/g/min and mean MPR was 2.20± 0.90 (table 3).

**Table 3:** CMR parameters based on metformin status. P value is considered significant at the <0.05 level and indicated by*. Continuous variables are presented as mean ± SD. Dichotomous variables are presented as number (%). **Adjusted for age, sex, LVEF, LV mass, LVEDV, hypertension, evidence of previous PCI, CABG, MI, BSA, ethnicity, CCB, nitrates, SGLT-2 inhibitors, GLP-1 agonists and gliclazide. MBF: Myocardial Blood Flow; MPR: MyocardialPperfusion Reserve; LV: Left Ventricle; LVEF: Left Ventricular Ejection Fraction; LVEDV: Left Ventricular End-Diastolic Volume; LGE: Late Gadolinium Enhancement, PCI: Percutaneous Coronary Intervention, CABG: Coronary Artery Bypass Graft surgery, MI: Myocardial Infarction, BSA: Body Surface Area, CCB: Calcium Channel Blocker; SGLT-2: Sodium glucose co-transporter 2 inhibitor; GLP-1-Glucagon like peptide agonist.

| CMR Parameter | Metformin (n=388) | No metformin (n=184) | P value |
| --- | --- | --- | --- |
| **LVEF (%)** | 59±14 | 53 ±18 | <0.001* |
| **LVEDV (ml)** | 149±50 | 174±71 | <0.001* |
| **LV mass (g)** | 117±34 | 127± 39 | 0.002* |
| **Global Stress MBF (ml/g/min)** | 1.74±0.6 | 1.62±0.5 | 0.019* |
| **Adjusted global stress MBF\*\* (ml/g/min)** | 1.74±0.03 | 1.67±0.04 | <0.0001* |
| **MPR** | 2.29±0.9 | 1.96±0.9 | <0.001* |
| **Adjusted MPR\*\*** | 2.27±0.04 | 2.03±0.07 | <0.0001* |
| **LGE ischaemic** | 115 (30) | 49 (27) | 0.502 |
| **LGE non-ischaemic** | 89 (23) | 50 (27) | 0.502 |
| **No LGE** | 183 (47) | 84 (46) | 0.502 |
| **1.5T scanner** | 127 (33) | 42 (23) | 0.015* |
| **3T scanner** | 261 (67) | 142 (77) | 0.015* |

Patients precribed metformin had significantly higher ejection fractions (59±14% vs 53±18%, P<0.001), stress MBF (1.74±0.6 ml/g/min vs 1.62±0.5 ml/g/min, P=0.019), MPR (2.29±0.9 vs 1.96±0.9, P<0.001) as well as significantly lower EDV and LV mass. There was no significant difference in the degree of scar burden between the two groups.

#### Healthy control group

All healthy volunteers were scanned on 3T scanners. The average stress MBF of the healthy control group was 2.05± 0.4 ml/g/min and average MPR was 3.40± 0.7.

##### Multivariable regression analysis

Multivariable linear regression was carried out to ascertain the extent to which metformin use is associated with stress MBF and MPR. The model was adjusted for factors known to be associated with differences in stress MBF and MPR including age, sex, LVEF, LV mass, LVEDV, hypertension, evidence of previous PCI, CABG surgery, MI, body surface area (BSA) and ethnicity as well as for medications including CCBs, nitrates, SGLT-2 inhibitors, GLP-1 agonists and gliclazide. Adjusted stress MBF and MPR values are reported in table 3. For stress MBF, the regression coefficient for metformin had a value of 0.05 ([0.04-0.17], P=0.21). This suggests that metformin use was associated with an increase of stress MBF by 0.05ml/g/min, although this did not meet statistical significance. For MPR, the regression coefficient for metformin had a value of 0.12 ([0.08-0.40], P=0.004). This suggests that the use of metformin is associated with an increase in MPR of 0.12, with statistical significance.

### Clinical outcomes

There were 82 first MACCE (14.3%) including a total of 27 (4.7%) deaths, 15 (2.6%) MIs, 18 (3.1%) strokes, 20 (3.5%) heart failure hospitalisations and 43 (7.5%) episodes of coronary revascularisation.

Table 4 shows first MACCE events by metformin status. Although there was no significant difference in first MACCE rates between the two groups, there was a significant reduction in number of deaths in the metformin group (9 (2.3%) vs 16 (8.7%), P<0.001). There was also a significantly lower incidence of stroke in the metformin group (8 (2.1%) vs 10 (5.4%), P=0.031).

**Table 4:** MACCE events by metformin status. P value is considered significant at the <0.05 level and indicated by*. Data represented as number of events with percentage in brackets.** First MACCE.

| **Event** | **Metformin**<br>**(n=388)** | **No Metformin**<br>**(n=184)** | **P value** | **All patients**<br>**(n=572)** |
| --- | --- | --- | --- | --- |
| **MACCE\*\*** | 53 (13.7) | 29 (15.8) | 0.503 | 82 (14) |
| **Death** | 9 (2.3) | 16 (8.7) | <0.001* | 25 (4) |
| **MI** | 12 (3.1) | 3 (1.6) | 0.307 | 15 (2.6) |
| **Stroke** | 8 (2.1) | 10 (5.4) | 0.031* | 18 (3.1) |
| **Heart Failure**<br>**hospitalisation** | 12 (3.1) | 8 (4.3) | 0.445 | 20 (3.5) |
| **Coronary**<br>**revascularisation** | 29 (7.5) | 14 (7.6) | 0.955 | 43 (7.5) |

Metformin was significantly associated with a reduction in all-cause mortality (figure 1). After adjusting for age and LVEF, the adjusted hazard ratio for all-cause mortality was 0.29 ([0.12-0.73], P=0.009 (table 5)). However, metformin use was not associated with reduced first MACCE rate after adjusting for age, LVEF, sex, LVEDV, LV mass, stress MBF, MPR and hypertension with an adjusted HR of 0.92 ([0.56-1.50], P=0.73) (table 5).

**Figure 1:**
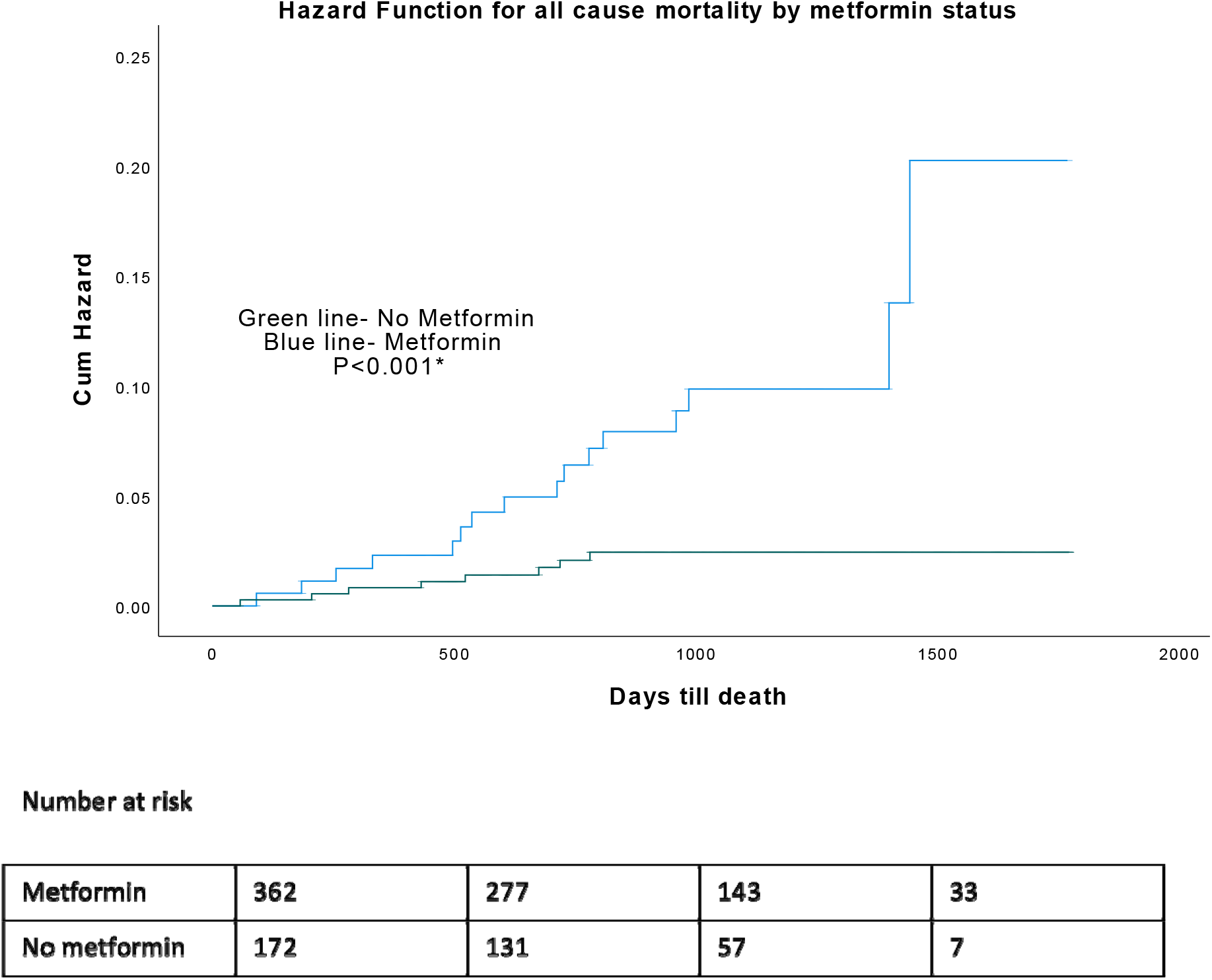
Kaplan-Meier hazard curves for all-cause mortality by metformin status. P value is considered significant at <0.05. Hazard curves are represented as blue line for no metformin and green line for metformin.

**Table 5:** Cox proportional hazard models. Death model was adjusted for age and LVEF. MACCE model was adjusted for age, sex, LVEF, LVEDV, LV mass, stress MBF, MPR, hypertension. Square brackets represent 95% confidence intervals. P value considered significant at <0.05 level and indicated by*. LVEF: Left Ventricular Ejection Fraction; LVEDV: Left Ventricular End Diastolic Volume; MBF: Myocardial Blood Flow; MPR: Myocardial Perfusion Reserve; MACCE: Major Adverse Cardiovascular and Cerebrovascular Event; HTN: Hypertension.

|  | **Death** | **MACCE** |
| --- | --- | --- |
| **Unadjusted** |  |  |
| Hazard ratio (Metformin) | 0.22 [0.10-0.52], P<0.001* | 0.87 [0.55-1.37], P=0.55 |
| **Adjusted** |  |  |
| Hazard ratio (Metformin) | 0.29 [0.12-0.73], P=0.009* | 0.92 [0.56-1.50], P=0.73 |

## Discussion

We sought to test the hypothesis that metformin use in patients with T2DM is associated with improved coronary microvascular function, improved survival and a reduction in MACCE.

The key findings in this study are that metformin use is associated with (1) improved MPR as a marker of CMD and (2) an improvement in all-cause mortality after adjusting for important parameters.

### Association between metformin and MPR

To the authors knowledge, this is the first clinical study that has demonstrated an association between metformin use and improved coronary microvascular function in patients with T2DM. Obesity, insulin resistance, and endothelial dysfunction closely co-exist throughout the natural history of T2DM^20^. Endothelial dysfunction is considered an important event in the development of microvascular complications in diabetes and is well recognised in patients with T2DM. Insulin resistance itself may be central to the pathogenesis of endothelial dysfunction. Patients with T2DM have been shown to have reduced MPR as manifestations of CMD caused by endothelial dysfunction^16, 17^. Metformin has been shown to improve endothelial function^21^. A previous study conducted in 16 asymptomatic patients recently diagnosed with T2DM using Positron Emission Tomography (PET), looked at the effect of metformin treatment alone or with a combination of glimepiride/metformin on coronary endothelial function^22^. It showed that at baseline, the response of T2DM patients to cold pressor test was reduced compared to controls, demonstrating endothelial dysfunction. Following treatment with a combination of metformin/glimepiride, myocardial flow reserve (MFR) and response to cold pressor testing MBF (% delta MBF) improved. Another study conducted in patients with T2DM showed an improvement in endothelium-dependent vasodilatation after administering metformin, assessed through forearm plethysmography. This change was also accompanied by a reduction of insulin resistance^11^. A study in rats who were fed a high fat diet showed an improvement in MPR after administering a 4-week treatment of metformin^23^. This study also showed that metformin treatment led to a robust adenosine-induced increase in myocardial perfusion (+81%). The authors postulated that metformin improved cardiac perfusion by restoring balance between vasoconstrictor and vasodilator effects of modest insulin resistance. We postulate that an improvement in endothelial function through the use of metformin in patients with T2DM, improves MPR. We speculate that this effect was not seen with stress MBF on multivariable regression analysis since MPR is a better measure of endothelial dysfunction^24^.

### Metformin and improved survival

Our data confirm previous reports that metformin is associated with improved overall survival. Despite having worse cardiovascular profiles, patients on metformin had higher ejection fractions compared to patients not prescribed metformin. We speculate that this may be secondary to more intense risk factor modification and drug treatment effects, for example with SGLT-2 inhibitors and GLP-1 agonists, although we cannot establish the cause from this study alone. The United Kingdom Prospective Diabetes Study (UKPDS) showed that metformin use reduced diabetes-related mortality by 42% (P=0.017), all-cause mortality by 36% (P=0.011), MI by 39% (P=0.01) and any diabetes related endpoint by 32% (P=0.002)^9^. In a recent systematic review, looking at 40 studies, metformin was shown to reduce cardiovascular mortality, all-cause mortality and cardiovascular events in CAD patients^25^. However, for MI and CAD patients without T2DM, metformin had no effect of reducing the incidence of cardiovascular events. In our study, metformin was not associated with an improvement in first MACCE rate. This is not surprising since patients prescribed metformin had higher cardiovascular risk compared to patients not prescribed metformin. In patients prescribed metformin there was also an increased prevalence of peripheral vascular disease and hypercholesterolaemia. The association between metformin use and improved survival is unclear; further research needs to be undertaken in this field. Recently, additional benefits of metformin have been reported including potential effects on cancer^26, 27^, cardiovascular disease^13^, liver disease^28^, obesity^29^, neurodegenerative disease^30^ and renal disease^31^.

### Improving mitochondrial function

T2DM is a metabolic disorder characterised by mitochondrial dysfunction and oxidative stress^32^. By increasing nitric oxide (NO) bioavailability, limiting interstitial fibrosis, reducing the deposition of advanced glycation end products (AGEs), and inhibiting myocardial cell apoptosis, metformin has been shown to reduce cardiac remodelling and hypertrophy, thereby preserving left ventricular systolic and diastolic function^33, 34^. Although we did not directly measure mitochondrial function in our study, these mechanisms may explain why patients taking metformin had higher ejection fractions, lower LVEDV and LV mass.

## Limitations

Several limitations should be acknowledged in this study. First, the study carries all the inherent limitations of an observational study and since subjects were non-randomised, the associations between metformin and improved survival and MPR found cannot imply causation and may be secondary to one or more confounders. One should also consider the potential for bias in the estimated coefficients due to the relatively small number of events and large number of co-variates. There is also the possibility that sicker patients may not have been prescribed metformin, leading to selection bias although this is unlikely to be the case in our cohort, since the patients prescribed metformin had more cardiovascular morbidity (increased prevalence of MI, PCI and CABG surgery, peripheral vascular disease and hypercholesterolaemia). It may also be plausible that patients prescribed metformin were better managed for conventional cardiovascular risk factors, due to the fact these patients tend to be higher risk patients. Furthermore, since events were documented using electronic patients records from different hospitals, there is a chance that events may have been missed. Patients on metformin in the current study were more likely to be on other medications such as gliclazides, nitrates, SGLT-2 inhibitors, GLP-1 agonists, and CCBs. Therefore, the improvement in MPR observed may have been mediated by one or more combination drug effects although we have adjusted for these in our multivariable linear regression analysis models and there is no evidence in the literature that these medications affect MPR. Furthermore, information on the duration of diabetes and the length of treatment with metformin was not collected in all patients. Despite these limitations, this is an indication that further research in this area, preferably in the form of a double blind, randomised prospective placebo-controlled clinical trial, would be able to address some of the points above.

## Conclusion

In patients with T2DM, we show that metformin use was associated with higher MPR, as a marker of microvascular function, and is associated with improved overall survival.

## Data Availability

The datasets used and analysed during the current study are available from the corresponding author on reasonable request.

## Abbreviations

CAD: Coronary Artery Disease
CMD: Coronary Microvascular Dysfunction
CMR: Cardiovascular Magnetic Resonance
T2DM: Type 2 Diabetes Mellitus
LV: Left Ventricle
LVEF: Left Ventricular Ejection Fraction
MI: Myocardial Infarction
MACCE: Major Adverse Cardiovascular and Cerebrovascular Events
MBF: Myocardial Blood Flow
MPR: Myocardial Perfusion Reserve

## Acknowledgements

The authors thank the clinical staff of the CMR department, and the National Institute of Health Research nurses based at Leeds General Infirmary.

## Author contributions

Study concepts/study design or data acquisition or data analysis/interpretation NS, KK, GSG, TK, LAEB, JLY, AP, RDA, ST, AC, EL, JM, GPM, MF, PK, JPG, PPS, SP; manuscript drafting or manuscript revision for important intellectual content: NS, SP, PPS, JPG,GPM,CG; approval of final version of submitted manuscript, all authors; agrees to ensure any questions related to the work are appropriately resolved, all authors; statistical analysis: NS, GSG, SP,TM,CG; and manuscript editing: NS,SP,JPG,GPM,CG.

## Ethics approval and consent to participate

All the patients provided written informed consent for their inclusion and approval was provided by the respective local ethics committees from each site and conducted in accordance with the Declaration of Helsinki.

Leeds data was obtained with ethical approval obtained by Leeds Research Ethics Committee (REC), Leeds, United Kingdom, reference 17/YH/0300, 18/YH/0168 and 18/YH/01900. Barts data used ethical approval from East of England, Cambridge Central Research Ethics Committee, United Kingdom, REC 14/EE/0007. Royal Free data was provided by the University College London/University College London Hospital Joint Committees on the Ethics of Human Research for recruitment at Royal Free Hospital (REC reference 07/H0715/101), United Kingdom. Leicester data ethical approval was provided by the UK Health Research Authority Research Ethics Committee (reference 17/WM/0192).

## Consent for publication

Consent for publication was obtained from all authors.

## Figure legends

*Central illustration: Metformin in patients with Type 2 Diabetes Mellitus (T2DM) associates with increased myocardial perfusion reserve (MPR) and is independently associated with increased overall survival. MBF: Myocardial Blood Flow; HR: Hazard Ratio; CI: 95% Confidence Intervals*

